# Are all underimmunized measles clusters equally critical?

**DOI:** 10.1101/2023.04.11.23288263

**Authors:** Sifat Afroj Moon, Achla Marathe, Anil Vullikanti

## Abstract

Disruptions in routine immunizations due to the COVID-19 pandemic have been a cause of significant concern for health organizations worldwide. This research develops a system science approach to examine the potential risk of geographical clustering of underimmunized individuals for an infectious disease like measles. We use an activity-based population network model and school immunization records to identify underimmunized clusters of zip codes in the Commonwealth of Virginia. Although Virginia has high vaccine coverage at the state level for measles, finer-scale investigation at the zip code level finds three statistically significant underimmunized clusters. To estimate the criticality of these clusters, a stochastic agent-based network epidemic model is used. Results show that different clusters can cause vastly different outbreaks in the region, depending on their size, location, and network characteristics. This research aims to understand why some underimmunized geographical clusters do not cause a large outbreak while others do. A detailed network analysis shows that it is not the average degree of the cluster or the percentage of underimmunized individuals in the cluster but the average eigenvector centrality of the cluster that is important in determining its potential risk.

## 1 Introduction

Measles is a highly contagious vaccine-preventable disease [Becker et al., 2016]. The United States (US) maintains a very high vaccination coverage level to induce herd immunity. However, disruptions in routine immunizations caused by the COVID-19 pandemic have become a major concern for the heightened risk of outbreaks of vaccine-preventable diseases, in general, and measles, in particular [Santoli, 2020, DeSilva et al., 2022, Murthy et al., 2021]. In 2021, twenty-five million kids were estimated to have missed their routine immunizations [Guglielmi, 2022].

According to a recent World Health Organization (WHO) report, measles cases were up by 79% worldwide in the first two months of 2022 compared to the same period in 2021. In 2022, many countries experienced measles outbreaks, such as India, Somalia, Yemen, Zimbabwe, and Pakistan. Zimbabwe’s Ministry of Health and Child Care reported more than 6,500 cases of measles on September 6, 2022 [Marima and Nolen, 2022].

In addition, spatial clustering of unvaccinated or under-vaccinated individuals may increase the risk of outbreaks. For instance, the 2019 measles outbreak in the US shows that spatial concentration of unvaccinated and under-vaccinated people can cause an epidemic outbreak even when the overall vaccine coverage rate in the region is high enough for herd immunity [Patel, 2019, Masters et al., 2020]. Several prior studies have demonstrated the existence of spatial underimmunized clusters of the Measles, Mumps, and Rubella (MMR) vaccine in the US with significantly lower immunization rates, using different methods [Cadena et al., 2019, Lieu et al., 2015, Dayan et al., 2005]. A scan statistics method is used by [Neill, 2012, Jung et al., 2010] to identify statistically significant geographical underimmunized clusters. This hypothesis testing approach for anomaly detection has previously been used in several studies to detect hotspots and anomalies in spatial datasets [Neill, 2009, Duczmal et al., 2006]. An extension of the scan statistics method for networks, used here in this research, discovers high-resolution clusters without any shape restrictions [Cadena et al., 2019].

However, not all underimmunized clusters pose an equal risk of outbreaks. For instance, although Virginia has a high immunization coverage rate of 95.8% among kindergartners [Seither et al., 2022], which is more than the target immunization rate for herd immunity, it has multiple underimmunized clusters, as shown in this paper. Further, Virginia has a high importation risk because of the two major international airports. For instance, a measles outbreak in 2021 affected the Central and Northern Health Regions of Virginia [Virginia Department of Health, 2021]. Additionally, as mentioned above, COVID-19 has disrupted routine immunizations, which might have led to a drop in MMR immunization rates across the state.

In this research, we formalize the outbreak risk of a cluster by its “criticality”, which is defined as the “probability of a large outbreak” caused by a single case of measles in the cluster. Our goal is to determine the criticality of different clusters. We focus on clusters that are most significant in terms of underimmunization rates. Given the limited public health resources, an objective measure of the criticality of clusters will help prioritize mitigation efforts.

Finding significant underimmunized clusters and computing their criticality is a challenging computational problem. We use a synthetic social contact network model for Virginia, and school-level immunization data in the state, along with a network scan statistics approach to find significant underimmunized clusters. We combine this with a detailed stochastic agent-based simulation framework to estimate the criticality of each significant cluster, by simulating outbreaks that originate in these clusters. We also examine how the criticality of clusters changes under a hypothetical 5% drop in MMR rate, possibly due to COVID-19. Finally, we study the demographic, geographic, and network factors associated with such clusters, which can help explain the potential risk of a cluster.

## 2 Methods

To find significant clusters and their criticality, we first identify statistically significant underimmunized clusters; for this purpose, we develop a zip code level spatial network from a detailed activity-based population contact network and use a network scan statistics method. Next, we investigate the criticality of each cluster by importing a single case of measles and simulating its spread using an agent-based model.

### 2.1 Synthetic social contact network model

We construct a contact graph *G*(*V, E*) for a population *V*, on which a disease can spread. In network *G*, nodes represent individuals, and edges represent a connection between two nodes. E.g., an edge (*u, v*) ∈ *E* between two people *u* and *v* is formed if they come into close proximity, which can enable disease spread. Additionally, we assume that each node *v* is associated with a geographical location, denoted by *loc*(*v*). The nodes in *G* have specific node properties, which come from geographic location and demographic characteristics such as age, gender, household size, household location, and household income. The edges are weighted, where weights are the contact time duration. This model combines various data sets from commercial and public sources, including the US census data, into a common architecture for creating a digital twin of the Virginia population. For more details on the construction of the social network, please see our earlier works [Eubank et al., 2004, Barrett et al., 2009, Thakur et al., 2022, Cadena et al., 2019].

The model places each individual in a household with others, and each household is located geographically in such a way that the aggregation of this synthetic population at the block group level is statistically equivalent to the US census block group data. The daily activities of individuals are assigned using time use surveys such as American Time Use Survey data [ATUS, 2021] and National Household Travel Survey Data [NHTS, 2021]. The geographic location for each activity is assigned from detailed land use data [HERE, 2021, NCES, 2021, LandScan, 2021, OpenStreetMap, 2021]. This activity-based network model also contains information about school locations, which are used for assigning schools to school-aged children [Cadena et al., 2019].

### 2.2 Finding significant underimmunized clusters

For this step, we first develop a zip code level spatial network, *G*_*z*_(*V*_*z*_, *E*_*z*_) for Virginia, where nodes are zip codes, and edges are connections between two zip codes. If two zip codes share a geographic boundary, they form an edge in the network *G*_*z*_. Each node or zip code *i* has two node properties: 1) population *P*_*i*_, and 2) immunization rate *r*_*i*_. We get *P*_*i*_ and *r*_*i*_ by aggregating the activity-based synthetic population network model *G* at the zip code level. Finally, we use a modified Kulldroff’s scan statistics method to find statistically significant underimmunized clusters in the network *G*_*z*_ [Cadena et al., 2019].

#### Immunization rate among children

We use the publicly available Virginia School Immunization Survey (SIS) record [SIS reporting, 2021] for Fall 2018 to calculate the MMR immunization rate among children (up to 17 years old). SIS contains immunization records for public schools. However, few kindergarten-level schools or schools with less than ten students report an overall vaccination rate instead of MMR specific immunization rate; and for some schools, there is no MMR immunization rate available in the SIS data. To handle the missing MMR immunization rate for these schools, we make the following assumptions:

1. If the MMR vaccination rate is not available for a school, but the overall immunization rate is available, then we use the overall immunization rate for that school.
2. In the synthetic population network model *G*, if the immunization rate for a school is not available from SIS data, we use the nearest neighboring school’s MMR immunization rate.

We find the nearest school by calculating the Haversine distance using the latitude and longitude. For children younger than eleven years old, we use the corresponding school’s kindergarten immunization record. To infer the immunization rate among kids 12-17 years old, we use the associated school’s 6th-grade immunization record. Previous studies have found that immunization status among children in the same household, even in the neighborhoods, schools, or jurisdictions, are positively correlated because of the geographic aggregation of vaccine refusal [Atwell et al., 2013, Lieu et al., 2015, Omer et al., 2008].

#### Immunization rate among adults

CDC’s ChildVaxView program reports state-level vaccination coverage among 19–35-month-olds via National Immunization Survey [ChildVaxView—CDC, 2021, Hill et al., 2018]. To obtain the state-level immunization rate among adults, we use a weighted average of the rates from the National Immunization survey report for 1995-2004 since the children from these years are adults in the current study year. We assign these immunization rates to the adults uniformly per age group.

#### Identify underimmunized clusters using network scan statistics

The network scan statis-tics, based on hypothesis testing, finds statistically significant underimmunized clusters or hotspots in network *G*_*z*_ [Kulldorff, 1997, Neill, 2009]. A cluster *C* ⊂ *G*_*z*_ in the proximity network *G*_*z*_ can have an arbitrary shape. We calculate the score function or scan statistic of a cluster of zip codes *C* as 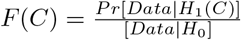 which is a likelihood ratio of the probability of the observed data (i.e., a certain level of underimmunization in *C*) generated under an alternative hypothesis *H*_1_(*C*), to the probability of the observations under the null hypothesis *H*_0_. We use the Poisson version of the Kulldorff scan statistic, which assumes that the observations are generated from a simple parametric distribution, Poisson distribution (a common assumption in epidemiological data analysis). The null hypothesis *H*_0_ is generated proportionally from the baseline count (1 − *μ*)*P*_*i*_, where *μ* is the state-wide immunization rate. The alternative hypothesis of a cluster *H*_1_(*C*) counts the vaccine distribution among nodes outside C; in *V*_*z*_ − *C*, the unvaccinated count comes from a rate proportional to the baseline counts. But, for the nodes within C, the counts are generated at a higher rate than expected. The objective is to find clusters that maximize the scan statistic *F* (*C*). We use the Monte Carlo sampling approach to compute the p-value for each cluster. Optimization over arbitrarily shaped clusters is computationally extensive as score function *F* (*C*) of interest is typically non-convex and NP-hard to optimize. We use a general dynamic programming method for optimizing a large class of parametric and non-parametric scan statistics [Cadena et al., 2017].

### 2.3 Criticality analysis

A stochastic agent-based network epidemic model allows us to investigate the impact of an underimmunized cluster.

#### Network epidemic model

We use an SEIR model for measles [Anderson and May, 1992, Keeling, 1997, Moon et al., 2019a, Moon and Scoglio, 2021], where an unvaccinated node can be in one of four health states: Susceptible (S), Exposed (E), Infected (I), and Recovered/Removed (R). Let **x**(*t*) be the health state vector at time t; *x*_*i*_(*t*) ∈ [0, 1, 2, 3] is the health state of an unvaccinated node *I* at time *t*. Here, 0, 1, 2, and 3 correspond to susceptible, exposed, infected, and recovered health state. A node *i* in network G has a vaccination status: vaccinated or unvaccinated. Let **v** be a *vaccination* vector: *v*_*i*_ ∈ [0, 1] denotes the probability that node *i* is vaccinated.

We assume that the vaccine has 100% efficacy (which is typically not true in practice); therefore, vaccinated nodes do not directly participate in the disease transmission. However, the presence of vaccinated nodes is important in the model as they affect the pathways to transmission by fragmenting the contact network.

We generally assume the source of the outbreak is a random infection in a subset *V*_*c*_ ⊂ *V*. Here, *V*_*c*_ is the set of nodes which are located in cluster *C*. We use *I*(**v**, *V*_*c*_) to denote the total number of infections given the vaccination vector **v** and source in *V*_*c*_. We use an agent-based stochastic individual network model to simulate the spread of measles in the contact network *G* (Fig. 2A). This model keeps track of disease progression and different health states of each node.

**Figure 1:**
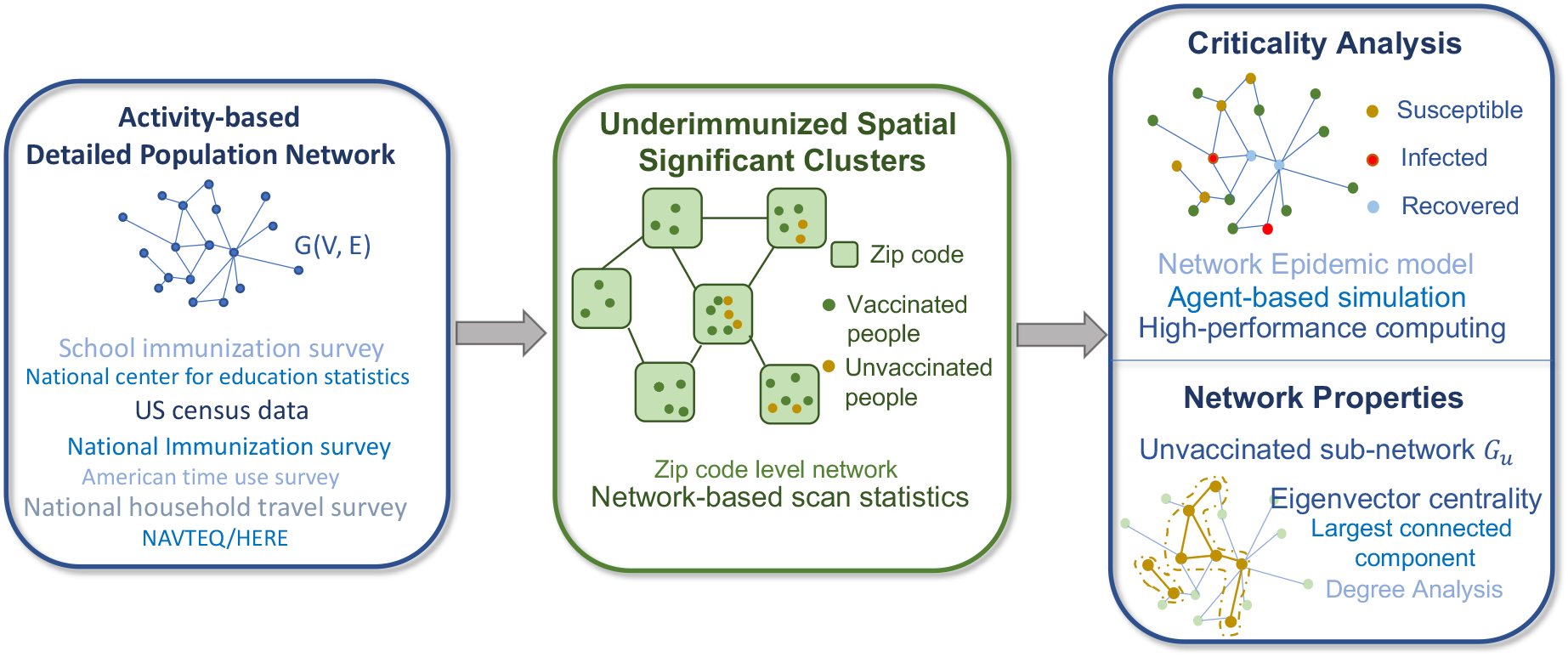
Four major components of the framework: 1) **An activity-based population network** *G*(*V, E*), where a node represents an individual, and an edge represents a contact between two people; 2) **Underimmunized spatial clusters** in a zip code level network *G*_*z*_(*V*_*z*_, *E*_*z*_), where nodes are zip codes, and a connection between two zip codes represents a geographically shared boundary; 3) **Criticality analysis** of each cluster using the stochastic network epidemic model; and 4) Understanding criticality by investigating **network properties** of unvaccinated sub-network *G*_*u*_.

**Figure 2:**
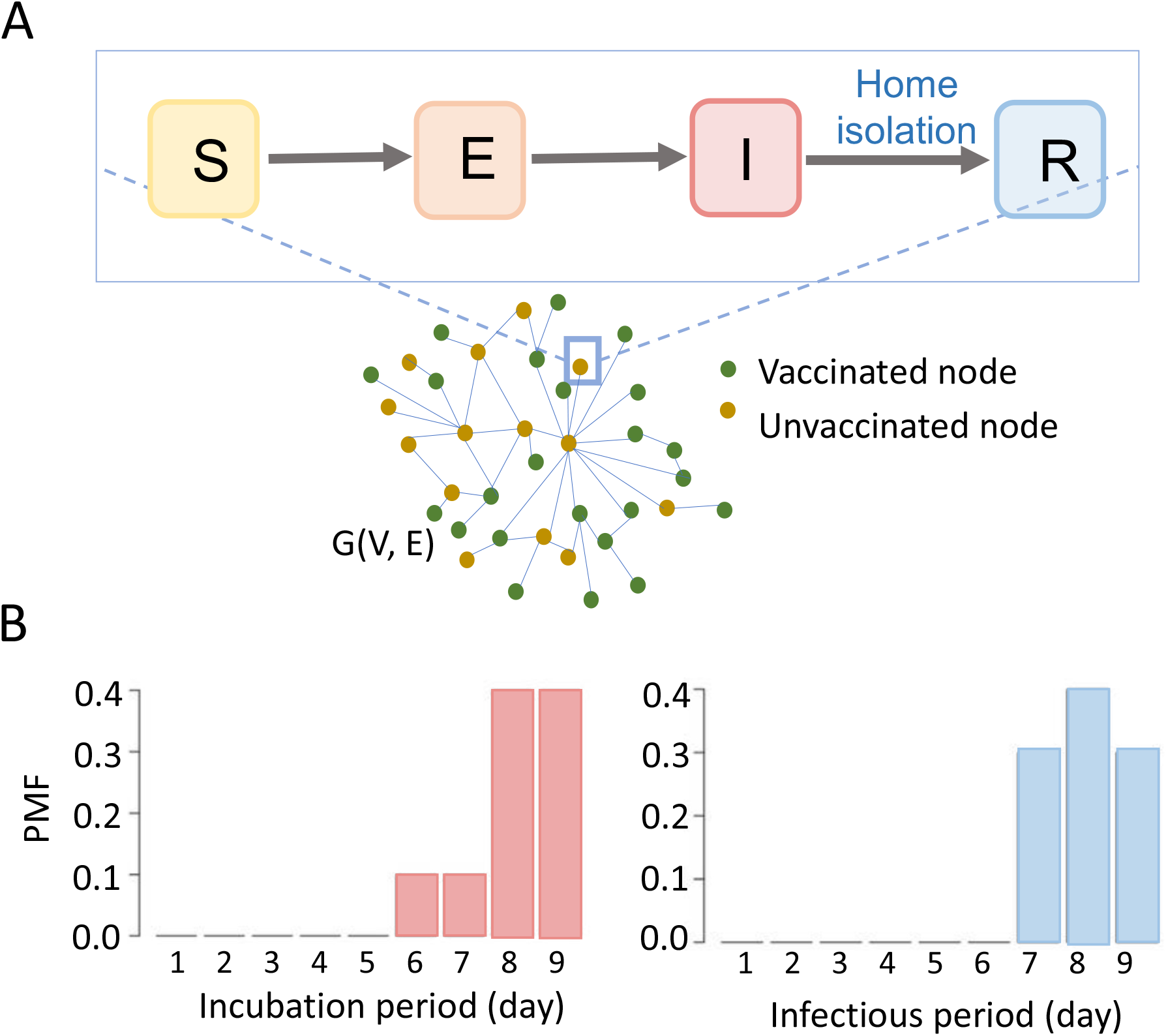
Disease transmission model. (A) Network SEIR (susceptible-exposed-infected-recovered) epidemic model, (B) probability mass function (PMF) for incubation period and infectious period.

Initially, we model that all the unvaccinated nodes are susceptible, and the disease starts from a random susceptible node. An infected node will transmit the disease to its susceptible neighbors in the contact network *G* with a transmission probability *β*. We use *β* = 0.5, as estimated from the recent New York City (NYC) outbreak that resulted in 649 cases between September 2018 and August 2019 [Thakur et al., 2022]. The outbreak size was calibrated for Virginia’s population size.

The disease model assumes that an exposed node will be infectious after a latent period. The maximum duration of the latent period is nine days and follows a discrete probability distribution {0, 0, 0, 0, 0, 0.1, 0.1, 0.4, 0.4}. The infectious or recovery period of an infected node is also nine days and follows a discrete probability distribution {0, 0, 0, 0, 0, 0, 0.3, 0.4, 0.3} (Fig. 2B) [Liu et al., 2015]. We assume that a recovered individual will not get reinfected.

#### Intervention

We assume that 90% (compliance rate) of the new infectious nodes will begin home-isolation after three days as the measles rash starts after 3-5 days [Liu et al., 2015]. In our network model, all non-home edges of the isolated cases are removed so they can only spread the disease to their household members.

#### Disease spread

An agent-based stochastic epidemic simulator *EpiHiper* [Machi et al., 2021, Eubank et al., 2010] simulates the spread of measles in the contact network *G*. It is a scalable system. Parallel programming and distributed memory systems allow it to handle millions of nodes and billions of edges. The simulation starts from a random infected node and runs for 365-time steps (or days). From 300 stochastic *EpiHiper* realizations, we compute the probability of having a large outbreak. If the number of infections exceeds *OC* = 500, we define it as a large outbreak. The total number of infections is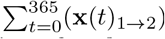; here, **x**(*t*)_1→2_ is the new infected nodes at day *t*.

#### Criticality of a cluster

We define the **criticality** of the set *V*_*c*_ ⊂ *V* in immunization **v**, denoted by crit(*V*_*c*_, **v**), as the probability of a large outbreak, when the initial infection occurs in subset *V*_*c*_. The probability of having a large outbreak is *Pr*[*I*(**v**, *V*_*c*_) *> OC*]; here, *OC* is the large outbreak condition. We consider a large outbreak if the total number of infections is more than *OC*.

### 2.4 Characterizing critical clusters

This research hypothesizes that all underimmunized clusters are not equally critical. When seeded, some clusters cause a large outbreak while others do not. To understand the potential reasons behind the criticality of clusters, we investigate their geographic and network properties. We explore the impact of geographic location, size, population density, and underimmunization rate on the criticality of a cluster. For network attributes, we measure their degree and eigenvector centrality. We also investigate the connected components.

#### Degree

The degree of a node *i* in a network is the number of neighbors of node *i*. If degree of a node *i* is *k*_*i*_, then average node degree of the network is 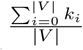. The average degree is important as it indicates the connectivity in a network [Barabási, 2013]. Moreover, high-degree nodes can act as hubs in spreading the disease [Dezső and Barabási, 2002].

#### Eigenvector centrality

It indicates the influence of a node in a network. The eigenvector centrality of a node is proportional to the sum of the centrality of its neighbors. It represents the spectral properties of the adjacency matrix *A* [Pastor-Satorras and Castellano, 2018]. In contrast to degree centrality, eigenvector centrality takes the entire network into account. This property makes eigenvector centrality particularly useful for understanding the influence of graph characteristics on epidemic spreading [Van Mieghem et al., 2008]. Eigenvector centrality *e*_*i*_ of a node *i* in a network *G*(*V, E*) is:

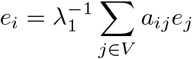

Here, *A* = [*a*_*ij*_] is the adjacency matrix of the network *G*, where *a*_*ij*_ = 1 if node *i* has a connection with node *j*, and *a*_*ij*_ = 0 otherwise. *λ*_1_ is the largest eigenvalue or spectral radius of the adjacency matrix *A* [Moon et al., 2019b]. The principal eigenvector, which corresponds to the largest eigenvalue, controls the structural and dynamical properties of a complex network. A large eigenvector centrality of a node indicates that it has many neighbors or important neighbors in the network. Eigenvector centrality is a good measure of a node’s spreading power [Canright and Engø-Monsen, 2006, Griffin and Nunn, 2012, Goltsev et al., 2012].

We use two networks for network analysis, namely, the full contact network *G*(*V, E*), and unvaccinated sub-networks *G*_*u*_(*V*_*u*_, *E*_*u*_) ⊂ *G*(*V, E*). In a sub-network *G*_*u*_, all the nodes *V*_*u*_ are unvaccinated; analysis of this sub-network is essential as the disease spreads only over this sub-network.

## 3 Results

### 3.1 Underimmunized clusters in Virginia

According to the CDC immunization record for the 2019–20 School year, MMR vaccine coverage among kindergartners in Virginia is around the target rate of 95% for herd immunity [Seither et al., 2021]. In our synthetic Virginia population network, the MMR immunization rate as estimated from various data sources, is 96.331% among children, and 91.496% for the overall population. We call the estimated immunization vector as the base immunization. The synthetic Virginia population contact network *G*(*V, E*) has 7, 688, 058 nodes and 371, 888, 622 edges; here, nodes are individual people. The zip code network *G*_*z*_(*V*_*z*_, *E*_*z*_) has 892 nodes and 2653 edges; here, a node represents a zip code. For each zip code, we calculate population and immunization rate by aggregating contact network *G*.

Although the MMR coverage is fairly high, the network-based scan statistics method finds three significant underimmunized clusters in Virginia (Figure 3). The vaccination rate in these regions varies from 88.2% to 90.6%. The statistical significance of these three clusters is measured from Monte Carlo Simulation: cluster 1 (log-likelihood score: 730.82, p-value: 9.99 * 10^−^4) is the most significant one, then cluster 2 (log-likelihood score: 141.01, p-value: 9.99 * 10^−^4) and cluster 3 (log-likelihood score: 98.10, p-value: 9.99 * 10^−^4).

**Figure 3:**
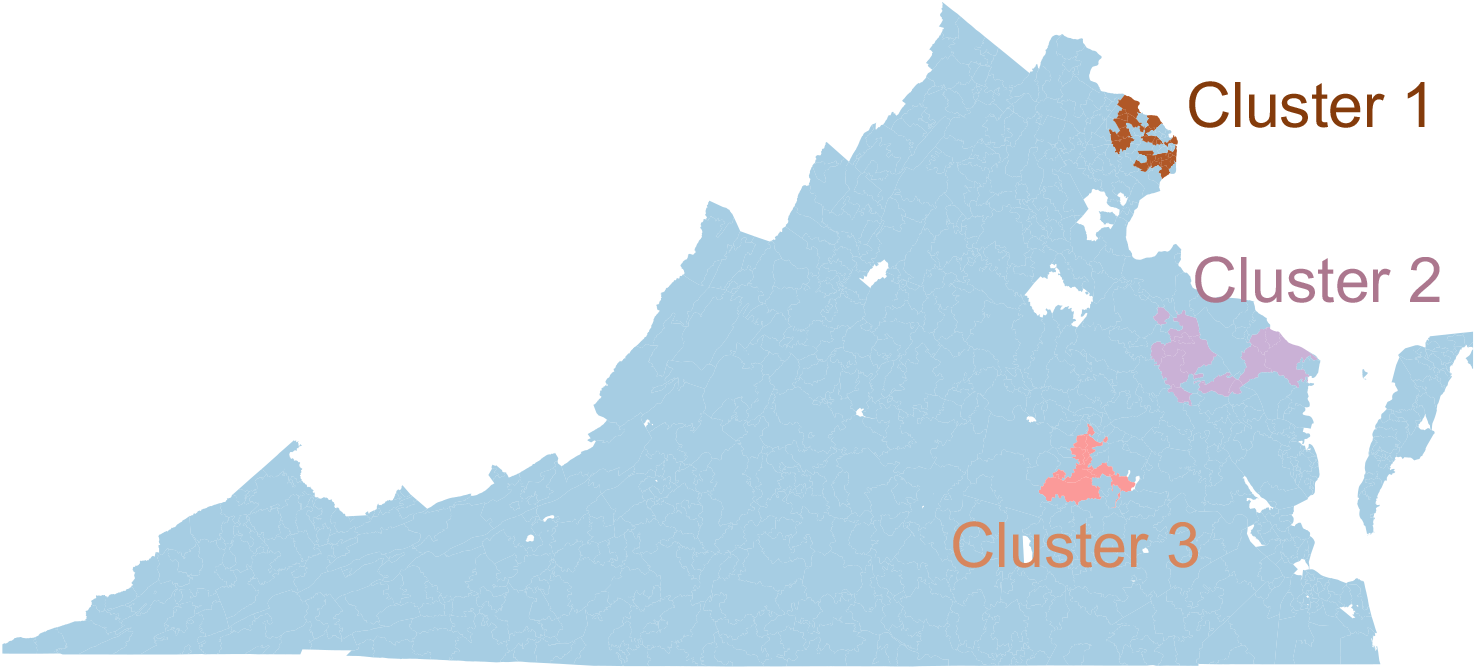
Critical underimmunized clusters in Virginia for MMR (measles, mumps, and rubella) vaccine.

Table 1 summarizes the geographic and network properties of the three clusters. The first cluster *C*_1_ is located in the highly populated part in northern Virginia. Population density per square mile in cluster 1 is 2684.2, which is over ten times the average population density per square mile in Virginia (≈ 202.6). This cluster contains 28 zip codes and is the largest underimmunized cluster in Virginia, with an immunization rate of 90.1%.

**Table 1:** Characteristics of the three statistically significant underimmunized clusters or hotspots in Virginia.

| Characteristic | Cluster 1 | Cluster 2 | Cluster 3 |
| --- | --- | --- | --- |
| Geographic location | northern Virginia | middle peninsula of Virginia | Richmond (capital) region of Virginia |
| No. of zipcodes | 28 | 13 | 9 |
| Population Count | 610464 | 24302 | 190832 |
| Underimmunized percentage | 9.9% | 11.8% | 9.4% |
| Population density per square mile | 2684.2 | 45.43 | 799.2 |
| Degree in $G$ | mean 47.9<br>median 42 | mean 33.9<br>median 28 | mean 40.8<br>median 36 |

Thirteen zip codes in the middle peninsula of Virginia form the second cluster *C*_2_. Although the population density in cluster 2 is only 45.43, which is less than one-fourth of Virginia’s average population density, its immunization rate is the lowest too, at only 88.2%. The second largest cluster: cluster 3, is located in the Richmond region of Virginia. The population density in cluster 3 is about four times higher than in Virginia.

These three clusters are unique concerning geographic location, shape, size, underimmunization rate, population density, and network characteristics. Even though the second cluster is the smallest and only affects a small population compared to the other two significant clusters, it can play a crucial role in disease spreading because of its position in the network and a low immunization rate.

### 3.2 Criticality of clusters

To calculate the criticality of a cluster *C*, we simulate an outbreak over the Virginia contact network *G*(*V, E*) using our agent-based simulation framework *EpiHiper*. The criticality of a cluster *C*_*i*_ is measured by estimating the probability of getting a large outbreak (similar to the NYC measles outbreak in 2019) if the epidemic starts from a random node in cluster *C*_*i*_. The seed is selected randomly from the unvaccinated age group of 5–17 years old in the cluster.

We apply four different seeding scenarios: 1) seeding randomly in Virginia (rand), 2) seeding in cluster *C*_1_, 3) seeding in cluster *C*_2_, and 4) seeding in cluster *C*_3_. The epidemic always starts from a single seed. For each scenario, we run 300 replicates and report the aggeregated result. The probability of a large outbreak for these four seeding scenarios is presented in Fig. 4. Our criticality analysis shows that the first cluster is the most critical, where seeding results in a large outbreak in 30% of the instances. In contrast, a random seeding in Virginia (rand) produces a large outbreak in less than 5% instances. Cluster 1 is the largest cluster and has the highest population density and node degree (Table 1).

**Figure 4:**
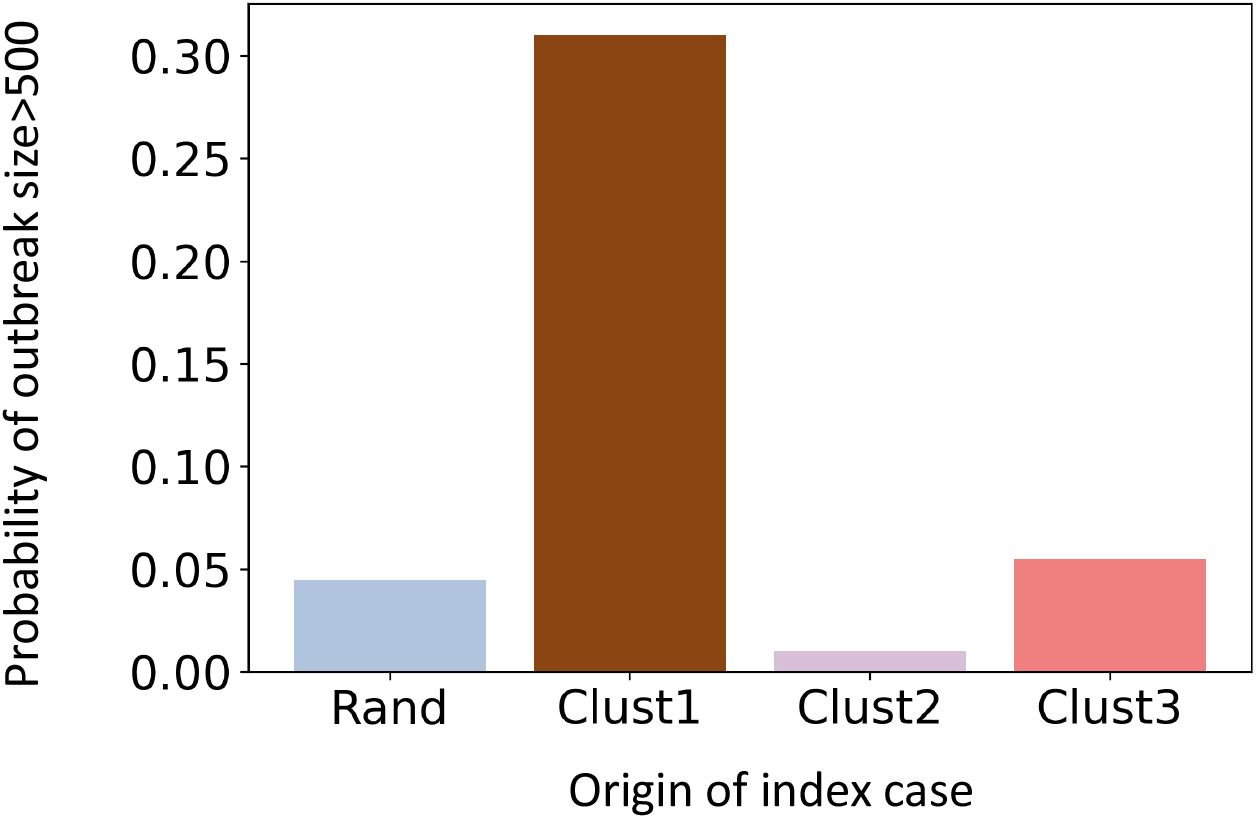
Probability of a large outbreak in Virginia in the base immunization for four different seeding scenarios: 1) random seeding in Virginia (Rand), 2) seeding in cluster *C*_1_ (Clust1), 3) seeding in cluster *C*_2_ (Clust2), and 4) seeding in cluster *C*_3_ (Clust3).

Fig. 4 shows that the criticality of cluster 2 is even lower than “rand”. The underimmunized pocket in cluster 2 does not increase the probability of large outbreaks, even though cluster 2 has the highest percentage of underimmunized individuals. Cluster 3 has a higher chance of causing an outbreak than “rand” but it is only marginally higher than “rand”, unlike cluster 1.

### 3.3 Risk analysis of drop in immunization rate

Due to COVID-19, routine childhood immunizations have been disrupted. To understand its impact on the criticality of the underimmunized clusters, we study a hypothetical scenario where a random uniform 5% drop in MMR immunization rate occurs among age group 12 years or younger in Virginia. The 5% drop in immunization rate among children (12 years or younger) in the State reduces the vaccine coverage in the network *G* by only 0.8%.

Fig. 5 presents the probability of a large outbreak for different seeding scenarios under the reduced immunization coverage. We find that the criticality of cluster 1 in the reduced immunization scenario *crit*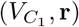 increases by 1.45 times its original criticality in the base case *crit*(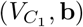. However, the criticality of the second cluster increases by 32.5 times of *crit*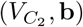 and of the third cluster increases by six times of *crit*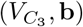. Only a 5% drop in immunization rate among kids (age 12 years or younger) makes the two benign clusters very critical. In the base case, the majority of the epidemic stays inside the cluster. For cluster 1, cluster 2, and cluster 3 seeding, the expected total incidence inside the cluster in base case is 67%, 77%, and 76% respectively (Table 2). In the 5% reduced immunization, the expected total incidence inside cluster for cluster 1, cluster 2, and cluster 3 seeding scenarios decrease to 37%, 3.6%, and 25.4% respectively, and increases the incidence outside the cluster. This is not surprising since many new nodes and connections are now part of the *G*_*u*_ network. However, the expected number of cases outside the cluster *C*_2_ go up significantly. For *C*_1_, *C*_2_ and *C*_3_, reduced immunization increases outside-cluster incidence by 40 times, 267 times and 131 times respectively. To understand possible underlying factors, we examine the network properties of the clusters.

**Table 2:** Size of the outbreak inside and outside clusters under base immunization and reduced immunization rates.

| Seeding | Base immunization | | 5% reduced immunization | | $\frac{5\% \text{ reduced immunization}}{\text{Base immunization}}$ | |
| --- | --- | --- | --- | --- | --- | --- |
|  | Inside cluster | Outside cluster | Inside cluster | Outside cluster | Inside cluster | Outside cluster |
| Cluster 1 | 1013.67 | 305.03 | 6987.99 | 12106.32 | 6.89 | 39.68 |
| Cluster 2 | 67.28 | 20.06 | 198.18 | 5357.66 | 2.95 | 267.0 |
| Cluster 3 | 60.88 | 19.11 | 854.48 | 2511.49 | 14.03 | 131.42 |

**Figure 5:**
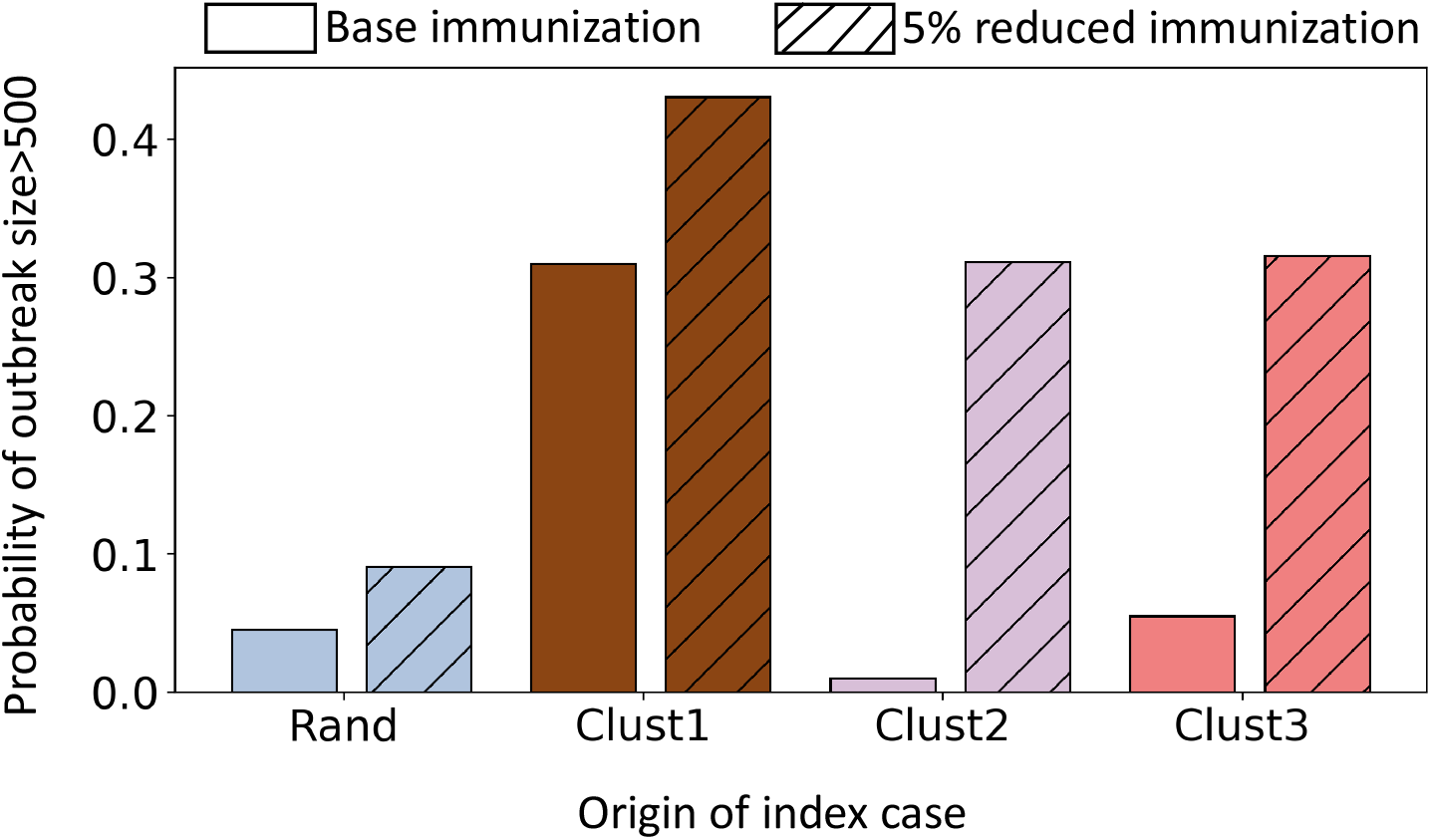
Probability of a large outbreak in Virginia under the base case and the reduced immunization case for the four different seeding scenarios.

### 3.4 Network analysis of the unvaccinated sub-network *G*_*u*_

In the base immunization case, the unvaccinated sub-network *G*_*u*_ has 653,811 nodes and 2,806,876 edges. The largest connected component has 525,586 nodes i.e. more than 80% of the nodes are connected. Table 3 shows that the average degree of clusters is not very different. However, the average eigenvector centrality in *C*_1_ is very high compared to *C*_2_ and *C*_3_, which explains why *C*_1_ has the highest criticality. Immunization drop increases the size of the unvaccinated network *G*_*u*_, connectivity, and average node degree. The average degree in *C*_2_ increases less than the other two clusters, i.e. only by 5.2%. The connections from *C*_2_ to outside nodes increase by only 7.5%, which is the lowest among all the clusters.

**Table 3:** Network Analysis of the unvaccinated sub-network *G*_*u*_ in the base immunization and reduced immunization.

| Network property | Base immunization | Reduced immunization | Increment |
| --- | --- | --- | --- |
| No. of nodes in $G_u$ | 653811 | 714164 | 9.2% |
| Largest connected component size in $G_u$ | 525586 | 588019 | 11.88% |
| No. of edges in $G_u$ | 2806876 | 3251182 | 11.87% |
| Average degree of the nodes in cluster $C_1$ in $G_u$ | 5.3141 | 5.6964 | 7.1% |
| Average degree of the nodes in cluster $C_2$ in $G_u$ | 4.829 | 5.0814 | 5.2% |
| Average degree of the nodes in cluster $C_3$ in $G_u$ | 4.096 | 4.4328 | 8.3% |
| Connections from the nodes in $C_1$ to outside in $G_u$ | 152840 | 167809 | 9.8% |
| Connections from the nodes in $C_2$ to outside in $G_u$ | 4905 | 5271 | 7.5% |
| Connections from the nodes in $C_3$ to outside in $G_u$ | 44358 | 48882 | 10.2% |
| Average eigenvector centrality of the nodes in cluster $C_1$ in $G_u$ | $5.4 * 10^{-4}$ | $5.68 * 10^{-4}$ | 5% |
| Average eigenvector centrality of the nodes in cluster $C_2$ in $G_u$ | $7.73 * 10^{-8}$ | $1.62 * 10^{-7}$ | 109.7% |
| Average eigenvector centrality of the nodes in cluster $C_3$ in $G_u$ | $6.43 * 10^{-8}$ | $7.96 * 10^{-8}$ | 23.6% |

Although an immunization drop adds a few new connections from cluster 2 to outside network but these new connections increase the average eigenvector centrality of *C*_2_ by 109.7%, which is significantly more than the other two clusters. It indicates that new connections increase the criticality of the cluster *C*_2_ by adding very important neighbors to nodes of *C*_2_.

To generalize the importance of eigenvector centrality on the criticality of a set of nodes, we investigate the SIR epidemic over the following two classes of random networks: 1) Barabási–Albert random network and 2) Erdős–Rényi random network. Barabási–Albert network is a popular model for designing social contact networks because of its scale-free power-law degree properties[Deville et al., 2016, Stephen and Toubia, 2009]. For both random networks, we choose 20, 000 nodes and around 601, 494 edges. We pick this relatively smaller network system than the Virginia contact network to experiment as they are computationally inexpensive. We consider random base vaccination coverage around 7.5%. In the unvaccinated sub-graph *G*_*u*_, we calculate degree and eigenvector centrality. We pick two sets of unvaccinated nodes in the sub-graph *G*_*u*_, with high and low eigenvector centrality but maintain the same number of nodes and average degrees in both sets.

We calculate the criticality of these two sets by using a stochastic SIR (susceptible-infected-recovered) compartmental epidemic model. For each set, we seed a random node and calculate the probability of a large outbreak. We consider an epidemic size greater than 100 to be a large outbreak for this smaller system. Our experiments find that the probability of a large epidemic is higher in the set with higher eigenvector centrality. Fig 6 presents experiment results from both random networks; here, the underimmunized rate is 7.5%. In both networks, the node in *G*_*u*_ is around 1500, and the average node degree is 4.5, which is close to the average degree of the unvaccinated sub-network of the Virginia contact network. Here, each set has 20 nodes. *β* and recovery rate in the SIR epidemic model is 0.3 and 1. In Fig 7, we also investigate other base underimmunized percentages and different levels of *β*.

**Figure 6:**
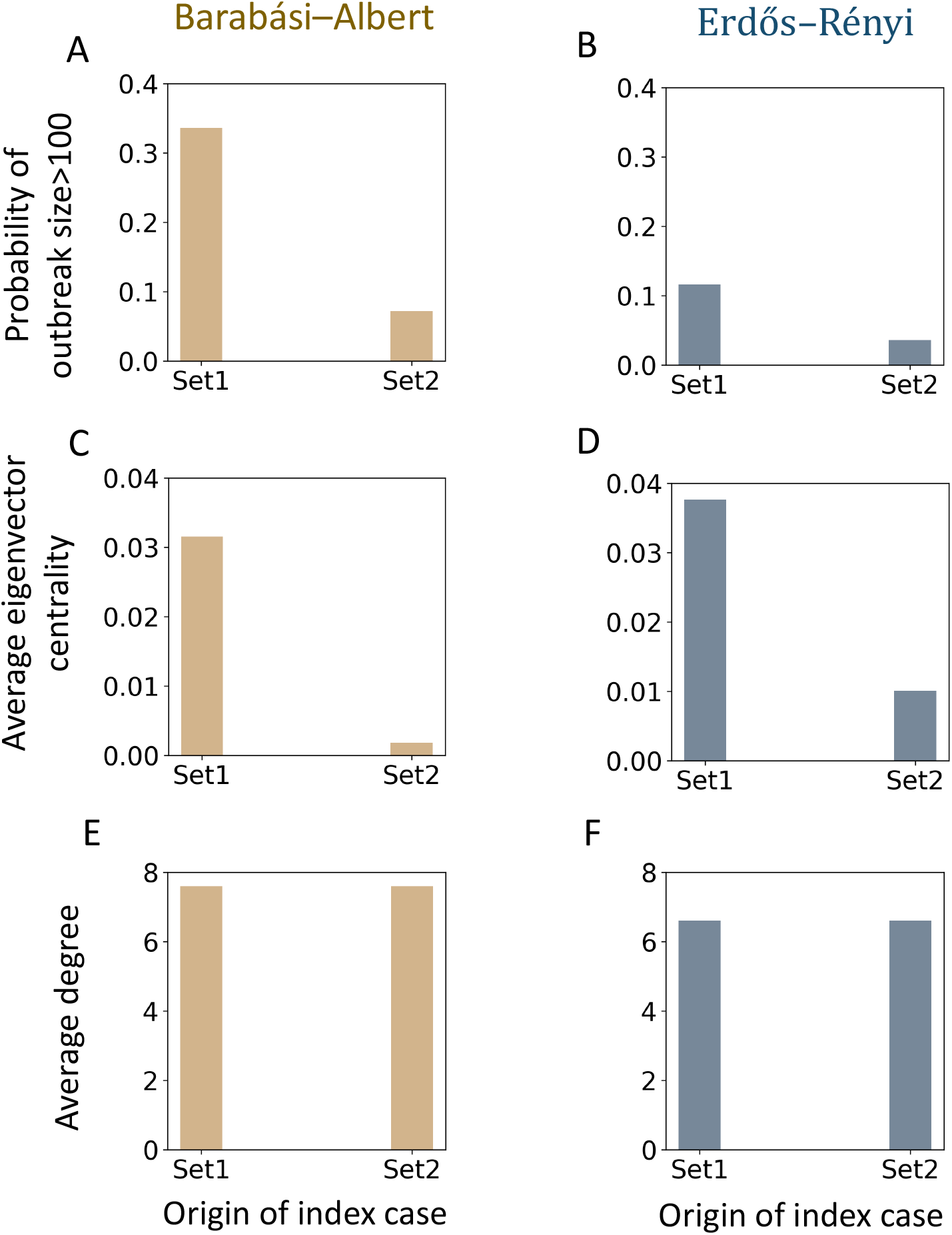
Criticality of two unvaccinated sets in the Barabási–Albert and Erdős–Rényi random networks; here, each set has 20 nodes, underimmunized rate is 7.5%, transmission rate, *β*, is 0.3, and recovery rate is 1. (A-B) probability of a large outbreak, (C-D) average eigenvector centrality, and (E-F) average degree.

**Figure 7:**
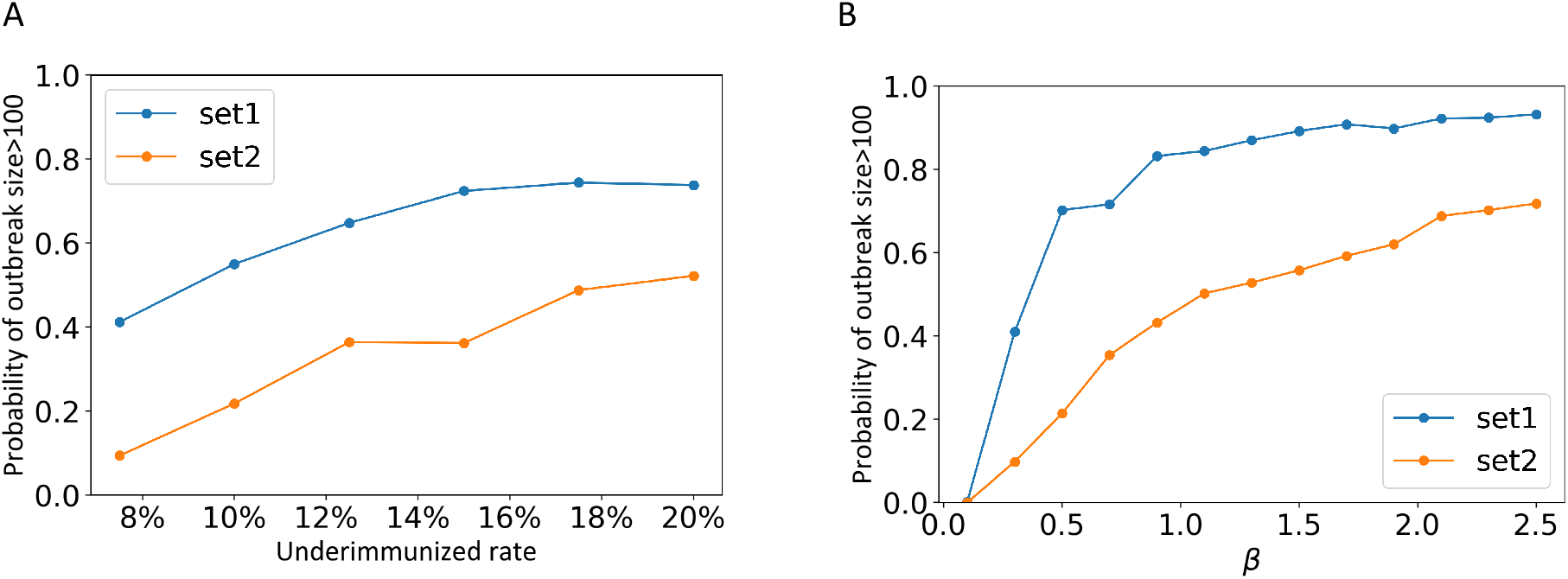
Sensitivity analysis of two unvaccinated sets in the Barabási–Albert random network. Both sets, set1 and set2 have 20 nodes and the same average degree, but set1 has higher eigenvector centrality.

## 4 Discussion

This research shows that all underimmunized clusters are not equal in terms of their risk of causing a large outbreak. Some are significantly more critical than others where even a small disruption in routine MMR immunization can disproportionately increase the risk of an outbreak. This can occur despite a very high immunization rate overall [Patel et al., 2019]. We use a system science model to find underimmunized spatial clusters in Virginia and measure their criticality. Our network-based Kulldorff’s spatial scan statistical method finds three significant irregular-shaped underimmunized clusters of zip codes in Virginia. These three clusters have very different geographic locations, sizes, population densities, immunization rates, and network properties (Table 1). The first cluster *C*_1_ is the largest one and is in a densely populated urban region. It also has a high average node degree and high average eigenvector centrality. The second cluster *C*_2_ is in a rural region. It is the smallest; however, it has the highest underimmunized percentage of individuals. The third cluster *C*_3_ is a large cluster, located near Richmond with a high population density.

To understand the criticality of a cluster, we use a stochastic individual-based network epidemic model that accounts for heterogeneous contacts and detailed immunization information. Our model considers vaccination status and home isolation as interventions at the individual level. The simulation results find that the criticality of underimmunized clusters are different. In the base case, the most critical cluster is the largest cluster *C*_1_. The probability of a large outbreak from seeding in cluster *C*_1_ is more than six times higher than the random seeding in Virginia. On the other hand, the criticality of the second significant cluster *C*_2_ is very low, even lower than the random seeding in Virginia. However, this benign cluster becomes very critical when MMR immunization rate drops by 5% among the age group of 12 years old or younger. Currently, this is a major concern since the COVID-19 pandemic has disrupted the routine immunization programs globally. According to CDC, more than 61 million doses of measles vaccine were postponed or missed because of the COVID-19 pandemic. In 2022, many countries around the world, including India, Yemen, and Somalia suffered measles outbreaks [Global Immunization, 2023].

We find that drop in immunization rate by 5% among children increases the criticality of all clusters but the percentage increase in criticality is significantly different for different clusters. For example, the criticality of the second cluster increases by 32.5 times the base immunization, after the drop. Reduced immunization rate also changes the incidence rates inside and outside of a cluster. In the 5% reduced immunization scenario, when *C*_2_ is seeded, the number of infections outside the *C*_2_ cluster is about 267 times the number of infections outside the cluster in the base immunization case. In contrast, the cases inside the cluster increase to only 2.95 times the base immunization case.

We find that the criticality of two sets of nodes can be very different if their eigenvector centrality is different, even though their average degree is similar. Previous literature has explored the importance of high eigenvector centrality in the context of spreading [Canright and Engø-Monsen, 2006, Griffin and Nunn, 2012, Goltsev et al., 2012] but this paper examines the criticality of a set of nodes in the context of vaccination and finds it to be an important characteristic in estimating the potential risk of clusters in the network.

## Data Availability

All data is publicly available

## 5 Acknowledgments

The authors would like to thank members of the Network Systems Science and Advanced Computing Division in the Biocomplexity Institute at the University of Virginia (UVA) for useful discussion and supports. They also would like to extend special thanks to Mandy Wilson and Jiangzhuo Chen for their valuable suggestions.

## 6 Additional information

This work is partially supported by National Institutes of Health (NIH) Grant R01GM109718, NSF (National Science Foundation) grants IIS-1955797, ACI-1443054, OAC-1916805, NSF Expeditions in Computing Grant CCF-1918656, CCF-1917819, US Centers for Disease Control and Prevention 75D30119C05935, DTRA (Defense Threat Reduction Agency) subcontract/ARA S-D00189-15-TO-01-UVA, and a collaborative seed grant from the UVA Global Infectious Disease Institute. The content is solely the responsibility of the authors and does not necessarily represent the official views of the sponsoring agencies.

## 7 Appendix 1

### 7.1 Sensitivity analysis of the random networks

We investigate the criticality of set1 and set2 in different base immunization levels (Figure 7A). Our result is robust for a wide range of base immunization (7.5% − 20%). We also consider various disease transmission probability *β*.

